# Planning Difficulties in Children and Adolescents with Hearing Loss across Development

**DOI:** 10.64898/2026.08.26.26361299

**Authors:** Katiac Monteseirín, Marta Méndez-Couz, Miguel Ángel Rivas-Fernández, Nélida M. Conejo

## Abstract

Children and adolescents with hearing loss frequently encounter reduced auditory access and delayed language development, factors that may influence the maturation of executive functions. This study examined developmental differences in planning, a core executive function, in 98 children and adolescents with hearing loss or normal hearing aged 7–18 years using the Tower of London task. Compared to normal hearing peers, participants with hearing loss made more unnecessary moves and rule violations and initiated problem-solving more rapidly, suggesting reduced preplanning efficiency and increased impulsivity. These group differences were most pronounced in adolescents, who showed faster initiation and greater movement inefficiency than age-matched normal hearing participants. Within the hearing loss group, adolescents displayed higher accuracy and longer initiation times than children, reflecting developmental improvements despite persistent gaps relative to hearing peers. Language development age did not alter the main effects. Findings indicate that reduced early auditory and language access may contribute to differences in planning development, highlighting the need for targeted executive functions support in educational and clinical settings for youth with hearing loss.

## INTRODUCTION

Executive functions (EFs) are a set of higher-order cognitive processes governed primarily by the prefrontal cortex (PFC) and its interactions with distributed brain networks (Friedman & Robbins, 2022; Menon & D’Esposito, 2022). These functions include inhibition, working memory, and cognitive flexibility (Baddeley, A. 2000; Miyake et al., 2000; Diamond, A., 2013; Dajani & Uddin, 2015), and extend to complex processes such as planning, emotional regulation, and problem-solving (Cristofori, Cohen-Zimerman, & Grafman, 2019; De Giacomo et al., 2021; Charry-Sánchez et al., 2022; Boerrigter et al., 2022; Lezak et al., 2012). Planning, in particular, requires the coordination of goal-directed behavior (Menon & D’Esposito, 2022), subgoal sequencing, and anticipatory decision-making. These processes rely on the maturation of PFC networks and their connectivity with sensory and association cortices (Benton, 1991; Eslinger, 1996; Barbey et al., 2012; Bruno et al., 2024).

The PFC undergoes prolonged maturation from childhood through adolescence, shaped by synaptic pruning, myelination, and increased functional connectivity with auditory and multimodal regions, including the superior temporal gyrus (Smieja et al., 2020). These neurodevelopmental changes underpin the gradual refinement of EFs across childhood and early adolescence (Anderson, 2002; Best & Miller, 2010). However, early auditory deprivation—such as in children with HL—may alter typical developmental trajectories of PFC–temporal lobe connectivity (Kronenberger et al., 2014; Pisoni et al., 2010). Structural and functional differences in deaf or hard-of-hearing children, including reduced white-matter integrity and altered activation patterns in fronto-temporal pathways (Kronenberger et al., 2018; Charry-Sánchez et al., 2022), support the view that auditory experience contributes to EF maturation. Both the auditory neurocognitive model (Kronenberger & Pisoni, 2020) and the language deprivation hypothesis (Hall et al., 2017) propose mechanisms through which HL may influence EF development. Reduced or inconsistent auditory access can limit auditory-driven cognitive scaffolding, whereas delayed language acquisition may restrict opportunities for verbally mediated planning and self-regulation. Thus, children with HL might exhibit differences in planning efficiency, impulse control, or problem-solving strategies relative to their typically hearing peers. Previous studies examining executive functioning in children with HL have reported difficulties across several domains, including working memory, inhibition, cognitive flexibility, and planning (Kronenberger et al., 2014, 2018; Charry-Sánchez et al., 2022). However, relatively few studies have specifically focused on planning using Tower-based paradigms. The Tower of London (TOL) task is particularly relevant in this context because it requires the integration of visuospatial working memory, inhibitory control, sequential organization, and anticipatory planning during goal-directed problem solving (Shallice, 1982; Luciana et al., 2009). Unlike broader EF measures, the TOL specifically captures planning efficiency and strategy formation under increasing cognitive load, making it especially sensitive to developmental changes in prefrontal functioning The current study examines how hearing loss may affect planning development across key developmental periods—childhood (7–12 years) and adolescence (13–18 years). Using the Tower of London (TOL) task, we investigated whether children (7-12-year-olds) and adolescents (13-18-year-olds) with HL differ from their typically developing peers in planning performance, and whether age and sex modulate these effects. We focused on planning because it is especially sensitive to frontal lobe dysfunction (Shallice, 1982) and provides insight into both behavioral control and neurodevelopmental maturity (Luciana et al., 2009).

We hypothesized that youth with HL would show reduced planning efficiency compared to NH peers, consistent with both auditory and language-based mechanisms of EF development. We also expected to observe developmental improvements in planning during adolescence, reflecting typical PFC maturation, and potential sex differences based on known disparities in EF and impulsivity across development (Rucklidge, 2008; Halpern, 2012).

## METHODS

### Participants

A total of 98 children and adolescents aged 7 to 18 years participated in the study (51 girls, 47 boys). Participants were recruited from mainstream educational settings in urban and suburban areas of Asturias, Spain. Specific ethnicity data were not collected. However, given the study’s location, the sample is likely predominantly of European descent. Two primary groups were defined based on hearing status: a control group with normal hearing (NH, n = 43) and a hearing loss group (HL, n = 55). Participation was voluntary and based on parental consent and participant assent. Consequently, the sample may reflect a degree of self-selection bias, particularly among families more willing to participate in cognitively demanding research protocols. Parental education level data were collected only for the clinical group (HL group) and categorized as: Primary education (equivalent to elementary school in the US educational system) (13.2%), Secondary education (equivalent to middle and early high school) (25%) and Higher education (college/university degrees) (57.4%). A fourth category, Unknown (4.4%), was included when data was not available. Data on parental education were not collected for the control group due to data availability. All participants primarily used oral language as their main mode of communication. Exclusion criteria included neurological, psychiatric, or developmental disorders; syndromic deafness; and any additional sensory or cognitive impairments.

Participants were also divided into two developmental subgroups based on sensitive periods of executive function (EF) development (Gariépy et al., 2019): children (7–12 years) and adolescents (13–18 years). In HL, n = 27 were children (n = 8 male, n = 19 female) and n = 28 adolescents (n = 14 male, n = 14 female). In the NH group, n = 18 were children (n = 9 male, n = 9 female) and n = 25 adolescents (n = 16 male, n = 9 female). This age stratification aligns with documented transitions in EF and frontal cortex maturation during early adolescence (Ciccia et al., 2009; Best & Miller, 2010).

### Hearing Status and Language Development

Participants in the hearing loss (HL) group used different forms of auditory assistance, including bimodal stimulation (cochlear implant + hearing aid; n = 7), cochlear implants (CI; n = 4), hearing aids (HA; n = 32), or no auditory assistance (n = 10). Device information was unavailable for two participants. These data are provided to characterize the heterogeneity of the HL sample. Verbal cognitive functioning was preliminarily assessed using the verbal subtest of the Kaufman Brief Intelligence Test (KBIT), in addition, a developmental language age was estimated for each participant using the Spanish version of the Peabody Picture Vocabulary Test (*Test de Vocabulario en Imágenes de Peabody, TVIP*), a widely used measure of receptive vocabulary and lexical comprehension (Monteseirín & Conejo, 2023). KBIT measure was used descriptively as a general indicator of verbal abilities and was not included in the primary statistical analyses. Raw scores were converted into age-equivalent scores using the normative tables provided in the test manual, yielding an estimate of each participant’s language developmental age. The gap between chronological age and language developmental age was computed to assess developmental language lag —a known moderator of EF performance (Hall et al., 2017; Kronenberger & Pisoni, 2020). Age-equivalent scores were selected because the primary aim was to quantify developmental delay relative to chronological age rather than normative language impairment. Although age-equivalent scores have recognized psychometric limitations, including reduced comparability across age ranges and non-normal distributions, they are commonly used in developmental research to characterize maturational discrepancies in pediatric populations (Maloney & Larrivee, 2007), including HL children (Ingvalson et al., 2023). Accordingly, language age-gap measures in the present study should be interpreted as developmental indicators rather than standardized estimates of language impairment.

### Executive Function Assessment

Executive function was assessed using the Tower of London (TOL) task (Shallice, 1982), a well-validated neuropsychological measure of planning and problem-solving. The version used comprised 10 levels of increasing complexity. Task instructions were provided orally, and each participant completed two practice trials before the testing began. Outcome variables included: TCS: Total Correct Score; TMS: Total Moves Score; TIT: Total Initiation Time (time before first move); TET: Total Execution Time (duration from first move to solution); TPST: Total Problem-Solving Time (TIT + TET); TRVS: Total Rule Violations (if the participant moves two beads at the same time); TTVS: Total Time Violations (tasks exceeding 2 minutes)

### Statistical analysis

#### Chronological age versus language development age

In order to evaluate group differences between the control group and the HL group in the gap between chronological age and the language development age, we conducted a univariate general linear model (GLM), including the group (two levels: control, hearing loss) as a fixed factor, the gap measure as dependent variable, and sex as covariate.

#### Age group

In order to evaluate the effect of age on cognitive performance in EF within the HL group, we conducted a multivariate GLM, including the age group (two levels: 7–12 years and 13–18 years) as fixed factor, with the TOL subscales as dependent variables. Additionally, sex and the gap between chronological age and language development age were included as covariates.

#### Control versus hearing loss group

To assess group differences in EF between control participants and the HL group, we conducted a multivariate GLM analysis with group (two levels: control, hearing loss) as a fixed factor. The TOL subscales were included as dependent variables, while sex and language development age were included as covariates. Additionally, to examine potential cognitive performance differences across developmental stages, we performed the same multivariate GLM analyses separately for two age groups (a subsample of participants aged 7–12 years and a subsample aged 13–18 years).

#### Age and sex-specific effects

To evaluate the effect of age and sex on cognitive performance in executive function within the NH and HL groups, we conducted a separate multivariate GLM per group. Each model included age group (two levels: 7-12 years, 13-18 years) and sex (female, male) as fixed factors, with the TOL subscales as dependent variables and language development age as a covariate.

All statistical analyses were corrected for multiple comparisons by using the False Discovery Rate Benjamini-Hochberg method (Benjamini & Hochberg, 1995).

Effect sizes for GLM analyses were calculated as partial eta squared (ηp²). Effect sizes are reported for all significant effects and interpreted using conventional thresholds (small = .01, medium = .06, large = .14).

#### Data Visualization

Group distributions for behavioral measures were visualized using kernel density– based violin plots with embedded box-and-whisker elements. The violins represent smoothed kernel density estimates (Gaussian kernel; bandwidth determined using Scott’s rule) extending beyond the observed data range to illustrate distributional shape. Embedded boxplots indicate the interquartile range (IQR), median, and whiskers corresponding to 1.5 × IQR.

For developmental and sex-specific analyses, data are presented as mean values with 95% confidence intervals (CI). All figures were generated using Python (Matplotlib and SciPy) and exported as vector graphics.

## RESULTS

### Group Differences (HL vs NH, all ages)

Children and adolescents with HL performed significantly worse on the TOL task compared to their NH peers. Specifically, the HL group made more unnecessary moves (TMS), more rule violations (TRVS), and initiated problem solving more rapidly (TIT) than NH participants (all FDR-adjusted *p* values ≤ .046; Table 1). These results suggest reduced preplanning and greater impulsivity in the HL group, consistent with previous reports of EF inefficiencies associated with auditory deprivation (Kronenberger et al., 2014, 2018; Charry-Sánchez et al., 2022). See Figure 1 and Table 1. No participants committed time violations, thus, the TTVS subscale was not included in the analyses.

**Figure 1.**
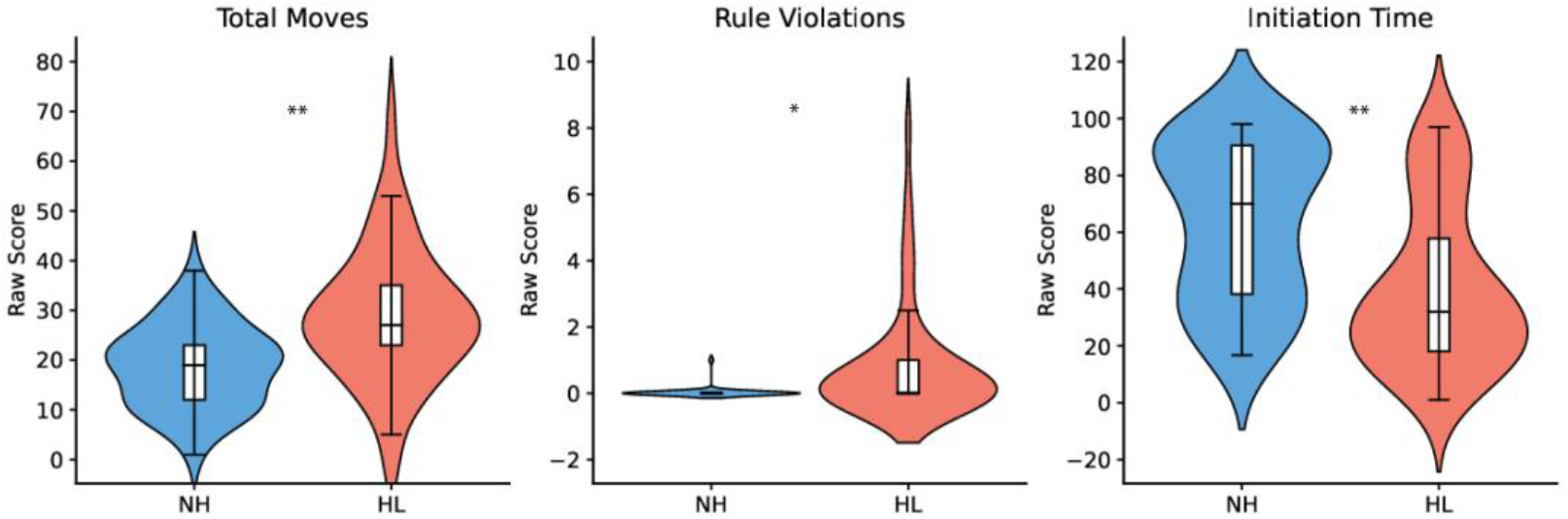
Group differences (NH vs HL, all ages). Group differences in Tower of London performance (NH vs HL, all ages). Violin plots with embedded boxplots show total moves, rule violations, and initiation time. Children and adolescents with HL displayed more unnecessary moves, more rule violations, and shorter initiation times than NH peers, reflecting increased impulsivity and reduced preplanning.* *p* < 0.05; ** *p* < 0.01.

**Table 1.**
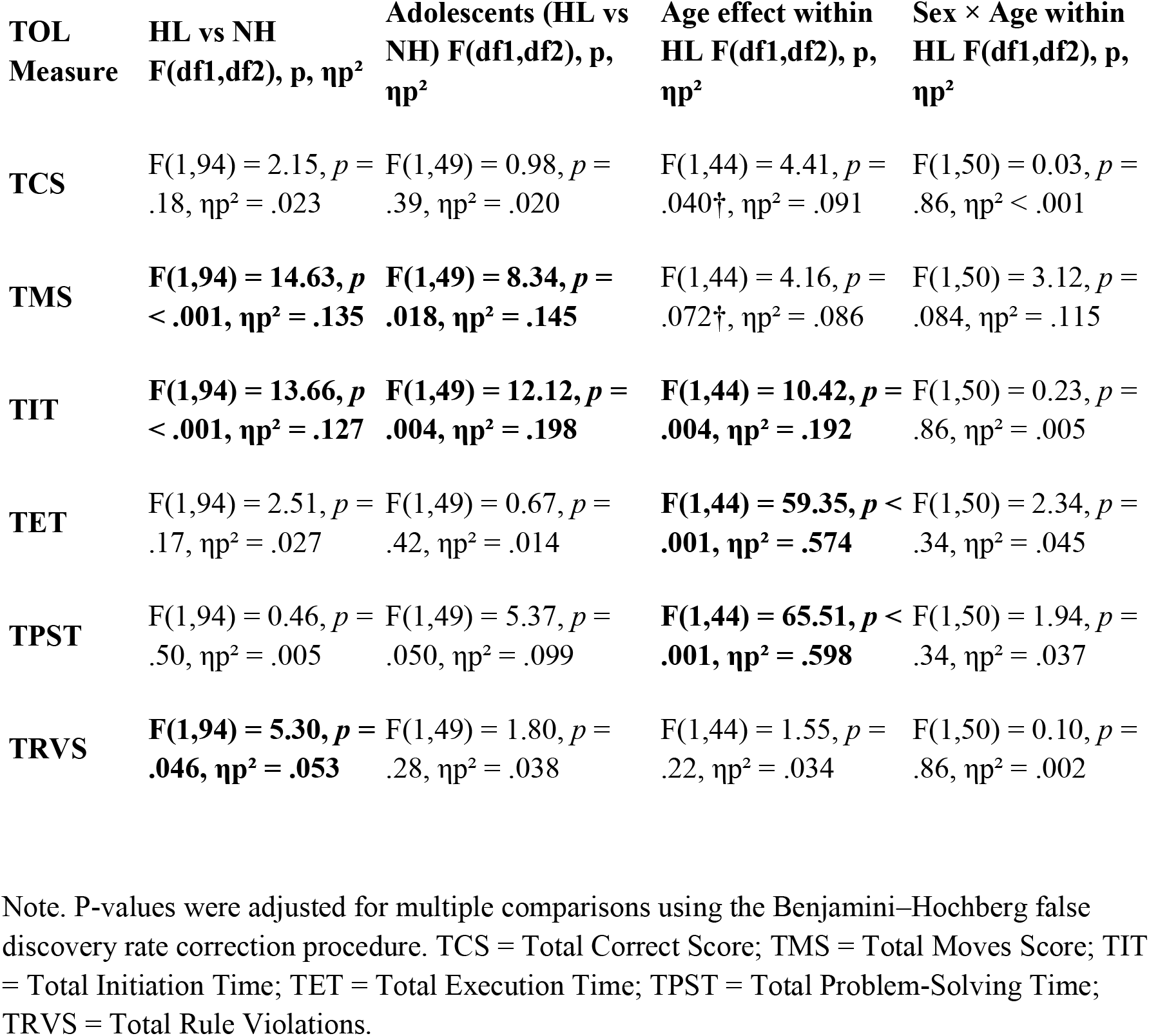
Summary of general linear model analyses for Tower of London (TOL) performance measures.

### Adolescent Subgroup Analyses

Differences between groups were particularly pronounced in adolescents (13–18 years). Compared to NH peers, adolescents with HL made significantly more moves (*TMS*: *p* < .01, ηp²= .145), displayed shorter initiation times (*TIT*: *p* = 0.001, ηp²= .198), and had reduced problem-solving times (*TPST*: *p* = .025, ηp²= .099). This pattern reflects more impulsive planning strategies during a developmental period when prefrontal cortex specialization and refinement are expected (Best & Miller, 2010; Johnson, 2011). See Figure 2 and Table 1.

**Figure 2.**
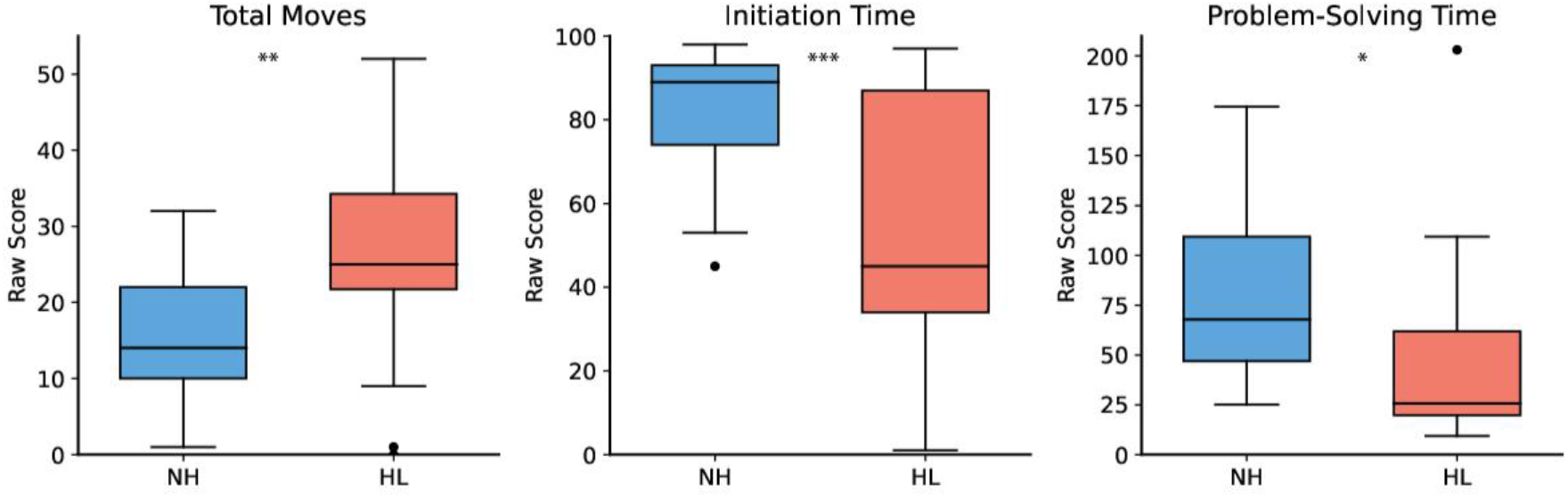
Planning in adolescents (NH vs HL). Planning performance in adolescents (13– 18 years, NH vs HL). Boxplots with jittered data points show that adolescents with HL made more total moves, initiated tasks faster, and had shorter problem-solving times than NH peers. These findings suggest increased cognitive impulsivity in adolescents with HL. * *p* < 0.05; ** *p* < 0.01; \*\*\**p* < 0.001.

### Age Effects within the HL Group

Within the HL group, developmental differences were observed. Adolescents (13–18 years) showed significantly higher total correct scores (TCS; *p* = .040), longer initiation times (TIT; *p* = .004), shorter execution times (TET; *p* < .001), and shorter total problem-solving times (TPST; *p* < .001) than children (7–12 years). Although children tended to make more unnecessary moves than adolescents, this difference did not remain significant after FDR correction (*p* = .072). These findings align with typical age-related improvements in EF efficiency (Anderson, 2002; Luciana et al., 2009), though overall performance remained below NH peers. See Figure 3 and Table 1.

**Figure 3.**
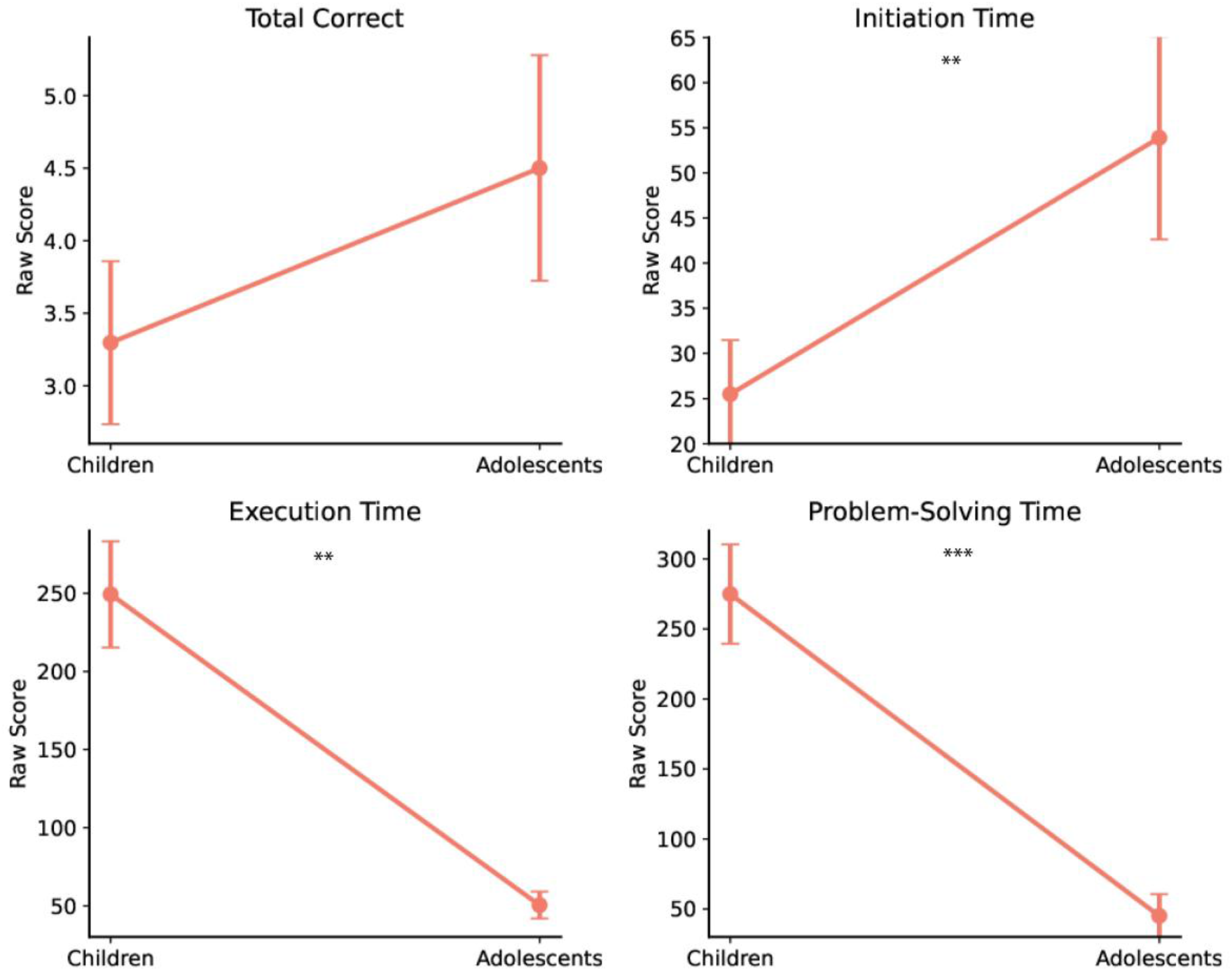
Developmental differences within HL group (Children vs Adolescents). Point plots show means and 95% CIs for total correct, initiation time, execution time, and problem-solving time. Adolescents with HL performed better than children on total correct and showed longer initiation times, consistent with maturation of planning strategies.\*\**p*<0.01; \*\*\**p*< 0.001.

### Sex × Age Interactions in HL

Exploratory analyses revealed a non-significant trend toward a sex × age interaction in unnecessary moves (TMS; p = .084, ηp² = .115). In particular, younger boys showed more unnecessary moves to complete the task than the younger girls (*TMS*: *p* < .02), consistent with impulsive responding. This trend is consistent with prior evidence for sex-related developmental differences in EF and spatial planning (Linn & Petersen, 1985; Rucklidge, 2008; Halpern, 2012). See Figure 4 and Table 1.

**Figure 4.**
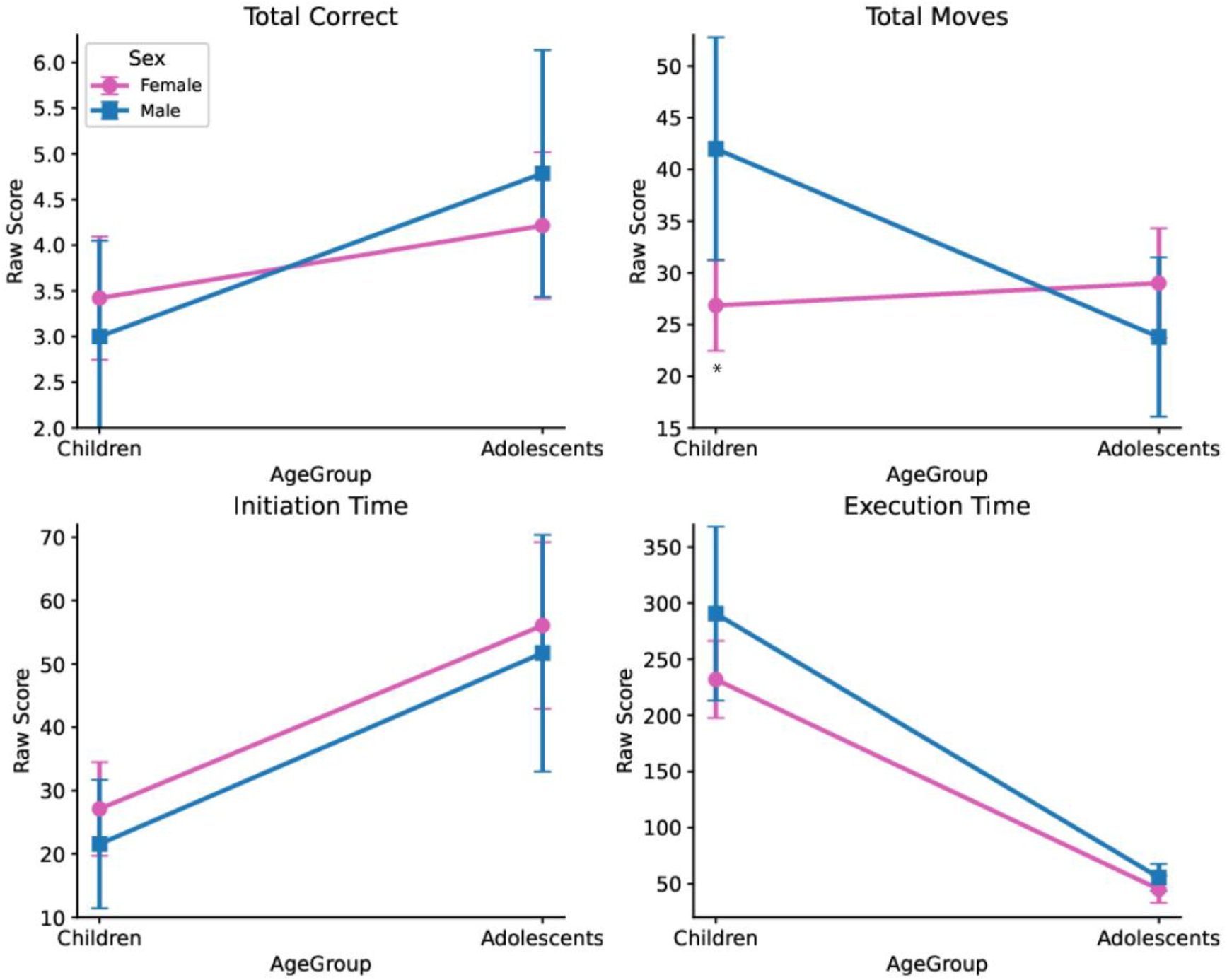
Sex x Age Interactions in the HL group. Sex × Age interactions in planning within the HL group. Interaction plots with means and 95% CIs show that adolescent boys outperformed girls on total correct and initiation time, while younger boys made more total moves and had longer execution times. These patterns highlight sex-specific developmental patterns of executive function in HL participants. \**p* < 0.05.

### Language Gap

Language gap (chronological age minus language age) was −4.67 ± 21.30 months in NH group and 12.62 ± 34.50 months in the HL group. It was included as a covariate and did not alter the main group effects, although consistent with previous work, poorer language proficiency was associated with lower EF performance (Hall et al., 2017; Monteseirín & Conejo, 2023).

## DISCUSSION

This study provides behavioral evidence that planning—a key executive function— develops atypically in children and adolescents with hearing loss (HL), particularly during adolescence. Using the Tower of London task, we found that participants with HL made more unnecessary moves and initiated tasks more quickly than their hearing peers, suggesting difficulties in preplanning and a tendency toward impulsive responses. These deficits were more pronounced in adolescents, aligning with neurodevelopmental models that emphasize the importance of adolescence for prefrontal cortex (PFC) specialization and refinement (Anderson, 2002; Best & Miller, 2010). Although the Tower of London (TOL) task is widely used to assess planning-related executive processes (Shallice, 1982; Luciana et al., 2009), it does not provide a comprehensive assessment of executive functioning (Diamond, 2013; Lezak et al., 2012). The TOL primarily captures visuospatial planning, sequential organization, working memory, and inhibitory control during goal-directed problem solving. Therefore, the present findings should be interpreted specifically as reflecting differences in planning-related executive processes rather than generalized executive dysfunction. In the context of hearing loss, these prefrontal maturation processes may depend on the quality and consistency of auditory and linguistic input, suggesting that reduced access to sound during early development could alter the emergence of efficient planning strategies.

The observed impulsivity in HL participants—reflected by shorter initiation times and more rule violations—suggests possible inefficiencies in top-down cognitive control processes. These behavioral patterns may mirror underlying neurodevelopmental alterations in fronto-temporal networks due to early auditory deprivation (Kronenberger et al., 2018; Smieja et al., 2020). Such deprivation can disrupt normative myelination, synaptic pruning, and PFC-auditory cortex connectivity, potentially delaying or altering the emergence of mature planning strategies (Pisoni et al., 2010; Charry-Sánchez et al., 2022). Thus, the observed planning behaviors in our sample may reflect the downstream effects of limited auditory-driven neural stimulation during early childhood. It is also important to acknowledge that differences in planning performance may reflect broader developmental, educational, or experiential factors associated with hearing loss rather than direct effects of auditory access alone.

Neuroimaging studies (e.g., fMRI, DTI) provide converging evidence that auditory deprivation disrupts normative connectivity between the prefrontal cortex and auditory cortices (Cardon & Sharma, 2013; Kral et al., 2016; Hribar et al., 2020). The dorsolateral PFC —a key hub for planning and goal-directed behavior—typically integrates auditory information from the superior temporal gyrus during planning tasks (Barbey et al., 2012). In children with early-onset deafness, studies have shown decreased white matter integrity (lower FA values) in fronto-temporal tracts such as the arcuate fasciculus and inferior fronto-occipital fasciculus (Li et al., 2012; Lyness et al., 2014), which correlates with poorer performance in EF-related tasks. Behavioral and neurodevelopmental studies also suggest atypical functional organization of executive-function networks in deaf and hard-of-hearing children (Kronenberger et al., 2018). Collectively, this supports a neurobiological mechanism where early auditory deprivation disrupts fronto-temporal connectivity, thus altering the developmental trajectory of prefrontal systems supporting EFs such as planning.

Exploratory analyses examining device type were not interpreted due to insufficient subgroup sizes, particularly for cochlear implant users. Our results might suggest that language exposure quality, timing of intervention, and broader environmental factors— such as family communication and cognitive stimulation—are critical determinants of developmental outcomes (Quittner et al., 2016; De Clerck et al., 2019). Our results indicate age-related improvements in planning among adolescents with HL, especially increased deliberation time and accuracy. This result may reflect partial catch-up in EF abilities, consistent with the “interactive specialization” theory of brain development, which posits increasing regional differentiation and network integration across adolescence (Johnson, 2011). However, persistent inefficiencies compared to NH peers suggest incomplete normalization, possibly due to compounded effects of early language delay (Monteseirín & Conejo, 2023).

Our exploratory sex-specific findings also support prior literature on sex differences in EF maturation: younger boys with HL showed more impulsive task behavior than their female counterparts, while in adolescence, boys show a descriptive tendency towards improved planning relative to girls. This aligns with the literature on earlier EF maturation in girls but greater visuospatial planning strengths in boys during adolescence (Linn & Petersen, 1985; Halpern, 2012).

From a translational perspective, these findings underscore the need for early, personalized interventions targeting planning and broader executive functioning. Neurocognitive training programs, particularly those adapted to the sensory profiles of children with HL, could leverage plasticity during adolescence to promote more efficient EF development. Moreover, integrating EF support into educational and psychosocial interventions may reduce risks of downstream academic, behavioral, and social-emotional difficulties (Holt et al., 2020; Hamed-Daher et al., 2024).

### Limitations and Future Directions

While this study offers valuable insights into planning-related executive function development in children and adolescents with hearing loss, several limitations should be noted. First, the sample size may limit the generalizability of some exploratory findings. Larger, more demographically diverse cohorts would strengthen the statistical power for detecting interaction effects. Second, although the Tower of London task is widely used to assess planning, it primarily captures visuospatial and sequential planning. Including additional EF measures, such as verbal planning or tasks assessing cognitive flexibility and inhibitory control, would provide a more comprehensive profile of executive functioning. Third, longitudinal studies are needed to determine whether observed differences represent delayed maturation or persistent atypical development. Finally, the lack of neuroimaging or electrophysiological data limits the ability to directly link behavioral performance with underlying neural mechanisms. Future research should integrate multimodal neurodevelopmental methods to elucidate the structural and functional correlates of EF difficulties in this population.

## CONCLUSION

This study provides behavioural evidence that planning difficulties in children and adolescents with hearing loss (HL) reflect age- and sex-specific variations in executive function (EF) development, likely underpinned by early reduced auditory and language access. Our findings reveal that, compared to typically hearing peers, youth with HL demonstrate shorter planning initiation times, more rule violations, and increased task impulsivity—behaviors that point to differences in the maturation and functional engagement of prefrontal executive systems.

Adolescents with HL showed some improvements over younger children, consistent with neurodevelopmental models of EF maturation; however, persistent deficits relative to controls suggest enduring alterations in prefrontal and fronto-temporal circuitry. The findings support both the auditory neurocognitive and language deprivation hypotheses and highlight the complex interplay between early sensory experience, language acquisition, and neurocognitive development.

Sex- and age-specific patterns suggest that EF development is not uniform in this population. These trends emphasize the importance of considering individual developmental profiles when designing interventions. Future longitudinal and neuroimaging studies are needed to better understand the neural basis of these observed behavioral differences.

These findings may support the inclusion of executive-function monitoring and planning-focused interventions within multidisciplinary follow-up programs for youth with hearing loss.

## Statements and declarations

### Ethical considerations

The studies involving humans were approved by the Vinjoy Foundation Ethics Committee. The studies were conducted in accordance with the local legislation and institutional requirements.

### Consent to participate

The participants and caregivers provided their written informed consent to participate in this study.

## Transparency and Openness

Due to privacy and ethical considerations involving minors, the raw datasets generated during the current study are not publicly available. De-identified data and analytical materials may be made available by the corresponding author upon reasonable request and in accordance with institutional confidentiality and ethical guidelines. The analyses reported in this study were not preregistered.

## Use of AI

The authors used artificial intelligence (AI)-assisted language tools to support editorial revision of the manuscript, including improvements in grammar, fluency, and clarity of expression. All scientific content, interpretations, analyses, and conclusions were developed and verified by the authors.

## Acknowledgements

We gratefully acknowledge the collaboration of Fundación Padre Vinjoy (Oviedo, Spain) and particularly its managing director, Adolfo Rivas, whose work in early intervention of children and adolescents with hearing loss made possible the participants’ recruitment.

